# Persistent gut microbiota alterations after ischemic stroke are modifiable by probiotic intervention

**DOI:** 10.64898/2026.10.15.26365107

**Authors:** Philip Melton, Adam Sorbie, Velma Aho, Christina Haslinger, Maili Zimmermann, Wiola Marut, Christian Jäger, Paul Wilmes, Corinne Benakis, Arthur Liesz

## Abstract

The gut-brain axis has emerged as a critical modulator in neurological diseases, including stroke. While acute post-stroke dysbiosis is well established, it remains unclear whether these alterations persist long-term and can be therapeutically modulated. This study addresses this gap through integrated metagenomic and metabolomic profiling in two independent clinical stroke cohorts, complemented by experimental models. Experimental stroke induced persistent changes in gut microbiota composition and altered short-chain fatty acid profiles. In patients, microbiota composition remained distinct from controls up to three months after ischemic stroke or transient ischemic attack (TIA), accompanied by persistent reductions in propionic and valeric acid and reduced abundance of *Prevotella*-associated microbial signatures. We performed a randomized, double-blind interventional trial (PRISE) testing a multispecies probiotic versus placebo, using household members as additional controls. Probiotic intervention led to microbiome compositional changes with increased abundance of beneficial taxa and significantly improved functional recovery. The tested probiotic strains showed anti-inflammatory and antioxidant effects in vitro. These findings suggest that targeted modulation of the microbiome represents a promising therapeutic avenue to improve chronic stroke recovery, warranting validation in larger Phase III clinical trials.

## INTRODUCTION

Stroke is a leading cause of mortality and disability.^1,2^ Previous studies have shown that ischemic stroke induces, along with numerous other comorbidities, gut microbiome dysbiosis and a subsequent disruption of the gut metabolome in the acute period.^3,4^ A possible explanation of this phenomenon lies in the transient intestinal paralysis that follows ischemic stroke.^5^ This post-stroke dysbiosis has been linked to intestinal immune alterations, systemic inflammation, neuroinflammatory responses, and functional outcome in experimental and clinical studies.^6,7,8^ However, the consequences of microbiome perturbation are context-dependent, as specific microbial configurations or experimentally induced dysbiosis can also attenuate post-stroke inflammation and improve outcome.^9^

A central challenge in microbiome research is to distinguish disease-relevant dysbiosis from physiological inter-individual variation, as gut microbiome composition varies substantially between individuals and is shaped by environmental, dietary, medication-related, and host factors.^10,11^ This is particularly relevant after stroke, where age, vascular risk factors, medication exposure, diet, acute illness, and hospitalization may all influence microbiome composition.^12,13^ Therefore, taxonomic profiling alone may be insufficient to define disease-relevant microbiome dysfunction. Microbial metabolites provide an additional functional readout of host-microbiome interactions.^14^ Among these, short-chain fatty acids, including acetate, propionate, butyrate, and valerate, are produced by bacterial fermentation of dietary fibers and regulate intestinal barrier function, immune cell differentiation, cytokine responses, and gut-brain communication.^15,16^

While acute post-stroke dysbiosis is well established,^17^ it remains unclear whether microbiome and metabolite alterations persist into the chronic recovery phase.^18^ It is also unknown whether these changes can be therapeutically modified. This is an important unresolved question, because sustained disruption of microbiome-derived immunomodulatory metabolites could affect post-stroke inflammation, tissue repair, and neurological recovery beyond the acute injury phase.

Here, we investigated microbiome composition and short-chain fatty acid profiles after experimental and clinical ischemic stroke, with a focus on persistent changes beyond the acute phase. We further tested whether probiotic intervention could modulate post-stroke dysbiosis in a prospective clinical trial and assessed the immunomodulatory effects of probiotic strains in vitro. Together, this approach aimed to define persistent microbiome-metabolite signatures after stroke and to explore their therapeutic potential.

## RESULTS

### Experimental stroke induces persistent gut microbiome and metabolome alterations

To investigate whether stroke induces sustained alterations of the gut microbiome, we subjected C57BL/6J mice to filament middle cerebral artery occlusion (fMCAo) and collected fecal samples immediately before surgery and on days 3 and 14 after stroke (Fig. 1A). Shotgun metagenomic profiling followed by NMDS based on Bray-Curtis dissimilarities revealed a significant time-dependent shift in microbiome composition (Fig. 1B). The relative abundances of individual bacterial species, such as *Bacteroidales species incertae sedis [meta_mOTU_v25_12701]* were perturbed and varied across all timepoints (Supp. Fig 1A), however alpha diversity as measured by oberved richness was not signifcantly different between the timepoints (Supp Fig 1B). We next assessed whether these compositional changes were accompanied by altered microbial metabolite profiles. Targeted mass spectrometry of ileal content at day 14 showed no significant difference in total short-chain fatty acid concentrations between naïve and fMCAo mice (Fig. 1C). However, ileal propionate was significantly reduced after fMCAo, while acetate showed a non-significant trend toward reduction (Fig. S1E). In plasma, total SCFA concentrations were reduced after fMCAo, although plasma propionate was increased (Fig. S1C,D). Thus, experimental stroke induced persistent changes in microbiome composition together with compartment-specific alterations in SCFA availability.

**Figure 1:**
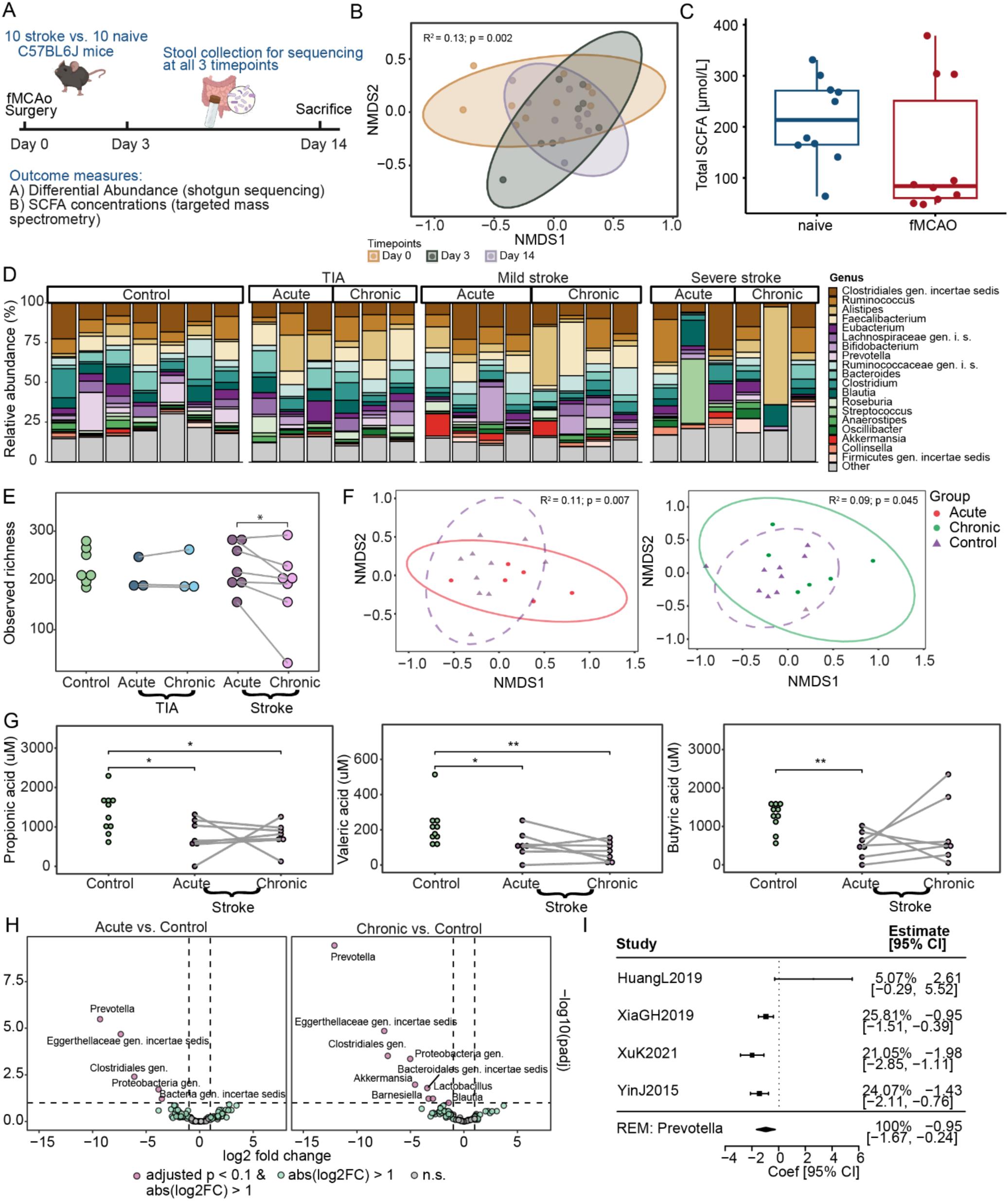
Persistent microbiome and SCFA alterations after experimental and clinical stroke (A) Experimental design. 8-week-old C57BL6J underwent filament middle cerebral artery occlusion (fMCAo) surgery. Stool was collected for shotgun sequencing before surgery, at 3 days post-fMCAo and 14 days post-fMCAo. Ileum content was collected for targeted mass spectrometry of the SCFAs at sacrifice (day 14). (B) Non-metric multidimensional scaling (NMDS) analysis using Bray-Curtis indices from sequenced stool samples (PERMANOVA; N = 8 - 10 per time point; R² = 0.13; p = 0.002) (C) Box-whisker plot of total SCFA concentrations measured in ileum content 14 days after fMCAo-operation or naïve animals (Wilcoxon; N = 10 per group; p = 0.19). (D) Bar graph of relative abundance (in %) of bacterial genera measured in stool of stroke/TIA patients of the SMB study or control patients from external dataset. (E) Dot plot of observed species richness measured between all patient groups (Wilcoxon; N = 3 - 7). (F) (left panel) NMDS analysis using Bray-Curtis indices between the control and acute stroke groups (Adonis; N = 6-7; R² = 0.11; p = 0.007), (right panel) NMDS analysis using Bray-Curtis indices between the control and chronic stroke groups (Adonis; N = 6-7; R² = 0.09; p = 0.045). (G) Dot plots of abundance (in µM) of propionic acid (left panel; paired t-test; N = 7 - 10), valeric acid (central panel; paired t-test; N = 7-10) and butyric acid (right panel; paired t-test; N = 7 - 10) in the control, acute and chronic stroke patient groups. (H) Volcano plots of differentially abundant bacterial genera (DESeq2; N = 6 - 7; Only taxa present in ≥ 10 samples were included). (I) Forest plot of Prevotella copri abundance in selected stroke cohorts. Shown are standardized mean differences, plot whiskers correspond to the 95% confidence intervals. (* denotes p ≤ 0.05; ** denotes p ≤ 0.005)

### The human gut microbiome remains altered three months after ischemic stroke

We next examined whether persistent microbiome alterations are also detectable after clinical ischemic stroke. Fecal and blood samples were collected from participants of the StrokeMicroBiomics cohort within 7 days and at 90 days after ischemic stroke or TIA and compared with age-and sex-matched control samples from the DeNoPa cohort (Fig. 1D; Table S1). Shotgun metagenomic sequencing showed a significant change in observed richness between the acute and chronic post-stroke time points, whereas richness did not differ significantly between stroke patients and controls and between time points in patients with TIA (Fig. 1E). In contrast, Bray-Curtis-based NMDS analysis demonstrated significant differences in beta diversity between controls and acute stroke samples, as well as between controls and chronic stroke samples (Fig. 1F). No significant beta diversity difference was observed between acute and chronic samples, suggesting that post-stroke microbiome composition remains distinct from controls at three months.

### Persistent SCFA alterations are associated with reduced Prevotella abundance after stroke

To assess microbiome function at the metabolite level, we quantified fecal SCFAs by targeted mass spectrometry. Total fecal SCFA concentrations were reduced in acute stroke compared with controls, but this difference was no longer significant at the chronic time point (Fig. S2B). However, propionic acid and valeric acid were significantly reduced both acutely and three months after stroke, whereas butyric acid was reduced acutely but not chronically (Fig. 1G). Untargeted plasma metabolomics further identified longitudinal changes in indole-3-propionic acid and pyruvic acid between the acute and chronic post-stroke time points (Fig. S2C).

Differential abundance analysis identified several genus-level features that were reduced at both acute and chronic time points compared with controls, including *Prevotella*, *Eggerthellaceae gen. incertae sedis*, *Clostridiales gen.*, and *Proteobacteria gen.* (Fig. 1H). Similar persistent alterations were observed at the mOTU level (Fig. S2A). *Prevotella* abundance correlated with fecal propionic, valeric, and butyric acid concentrations across all samples, and with propionic and valeric acid in stroke-only analyses (Fig. S3). To assess whether the reduction in *Prevotella* observed in our cohort was consistent with previous clinical stroke microbiome studies, we performed a post-hoc analysis of published 16S rRNA gene amplicon sequencing datasets reporting altered Prevotella abundance after stroke. Across four independent studies, effect estimates were directionally consistent with lower *Prevotella* abundance in stroke cohorts compared with controls, although the magnitude of the effect varied between studies (Fig. 1I). A random-effects summary estimate confirmed an overall reduction of *Prevotella* in stroke. Thus, the persistent *Prevotella* depletion observed in the SMB cohort is supported by prior clinical stroke microbiome data and may represent a reproducible feature of post-stroke dysbiosis. These findings link persistent taxonomic alterations after stroke to reduced SCFA concentrations, although the correlative nature of the analysis does not establish direct metabolite production by these taxa.

### Probiotic intervention modulates post-stroke microbiota and is associated with improved functional recovery

To test whether post-stroke dysbiosis can be therapeutically modified, we conducted a randomized, double-blind prospective clinical trial in patients with ischemic stroke. Participants were enrolled within 7 days after stroke onset and randomized to twice-daily probiotic or placebo treatment for 90 days, after which stool samples were collected for shotgun metagenomic sequencing. Household members provided stool samples at the same time point and served as a control group (Fig. 2A).

**Figure 2.**
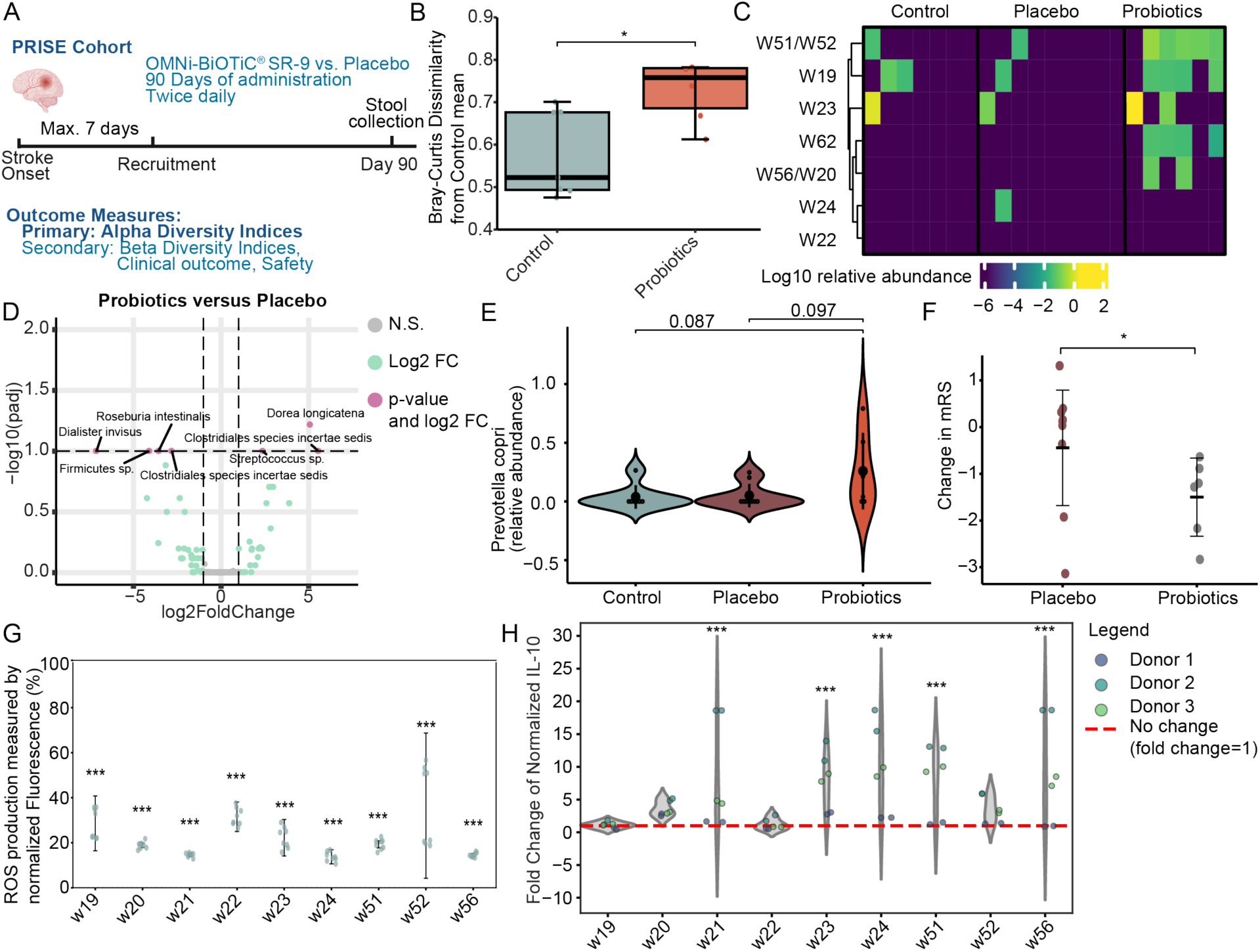
Probiotic intervention modulates post-stroke microbiota composition and improves functional recovery. (A) Study design (PRISE). Stroke patients were recruited within 7 days of symptom onset and randomized to treatment with OMNi-BiOTiC^®^ SR-9 or placebo. Study participants received intervention for 90 days, stool samples were then collected for shotgun sequencing. Household members of participants were recruited as a control group and donated stool samples simultaneously to the treatment groups. A total of 22 participants were recruited, comprising 6 receiving probiotic intervention, 9 receiving placebo intervention and 7 household controls (B) Box-whisker plot of Bray-Curtis dissimilarity from the household control mean. (Wilcoxon; N = 6 - 7; p = 0.035). (C) Heatmap of relative abundance (scale: Log^10^) of bacterial species contained within OMNi-BiOTiC^®^ SR-9 across the participant groups. Columns represent individual participants. (D) Volcano plot of differential abundance of mOTUs between probiotic and placebo treated groups (DESeq2; N = 6-9). (E) Violin plot of relative abundance of Prevotella copri in participant groups (t-test; N = 6 - 9). (F) Dot-whisker plot of change in mRS between baseline and 3 months in the treatment groups (Wilcoxon; N = 6 - 9; p = 0.042). (G) Scatterplot of RAW 264.7 cell cultures stimulated with 200 µm tertiary butyl hydroperoxide and 5% supernatant from a bacterial strain. Radical oxygen species levels were determined via DCF fluorescence at excitation/emission wavelengths of 488 nm/525 nm. The relative changes in fluorescence were normalized to RAW 264.7 cells only stimulated with 200 µm TBHP. (Linear mixed effects model; N = 8 per group) (H) Violin plot of fluorescence intensity fold changes of IL-10 in supernatant of human PBMCs from 3 donors cocultured with probiotic strains. PBMCs were cocultured 1:1 with 9 different bacterial strains and stimulated with Anti-CD3 and Anti-CD28 antibodies for 24 hours before Luminex measurement of IL-10. PMBCs stimulated without co-culture were used to normalize the measurements on a per-donor basis (Linear mixed effects model; N = 6 per group). (* denotes p ≤ 0.05; ** denotes p ≤ 0.005; *** denotes p ≤ 0.0005)

Global microbiome composition did not differ significantly between probiotic-treated patients, placebo-treated patients, and household controls by Bray-Curtis-based ordination analysis (Fig. S4C). Similarly, the prespecified alpha-diversity endpoint was not significantly different between groups at 90 days (Fig. S4D). However, probiotic-treated patients showed a significant mean Bray-Curtis dissimilarity from household control microbiome profiles, suggesting a partial modulation of the microbiome in probiotic-treated patients (Fig. 2B).

We next assessed whether probiotic administered species could be detected in recipients. Species-level analysis showed enrichment of probiotic species in stool samples from the probiotic group compared with placebo-treated patients and household controls, indicating treatment-associated persistence of administered species at 90 days (Fig. 2C). Differential abundance analysis further identified taxonomic shifts between probiotic- and placebo-treated patients, including higher abundance of *Dorea longicatena*, Clostridiales species incertae sedis, and *Streptococcus* sp., and lower abundance of *Roseburia intestinalis*, Firmicutes sp., Clostridiales species incertae sedis, and *Dialister invisus* in the probiotic group (Fig. 2D). Among taxa identified as persistently reduced after stroke in the SMB cohort, *Prevotella copri* showed a trend toward increased abundance in probiotic-treated patients, however these changes were only observed in a few individuals (Fig. 2E). In addition, Eggerthellaceae gen. incertae sedis was increased in the probiotic group, whereas *Barnesiella* was reduced in placebo-treated patients compared with household controls (Fig. S4G,H). Thus, probiotic intervention did not broadly restore microbiome diversity, but induced measurable, treatment-associated changes in microbiota composition.

We then evaluated clinical recovery from baseline to 90 days. Patients receiving probiotic treatment showed a significantly greater reduction in modified Rankin Scale scores compared with placebo-treated patients, indicating improved global functional recovery (Fig. 2F). No significant between-group differences were observed in 90-day MoCA scores or in NIHSS change from baseline to follow-up (Fig. S4I,J). Together, these findings indicate that probiotic intervention partially modulates post-stroke microbiota composition and is associated with improved functional outcome in this prospective randomized trial.

### Probiotic strains reduce oxidative stress and modulate PBMC cytokine responses in vitro

To assess whether the probiotic strains used for the clinical intervention may exert immunomodulatory effects, we performed two complementary in vitro assays. First, RAW 264.7 macrophages were pre-treated with supernatants from the individual probiotic strains and then exposed to tert-butyl hydroperoxide to induce oxidative stress. Pre-treatment with 5% probiotic supernatant reduced ROS production across all tested strains compared with stimulated control cultures (Fig. 2G). This effect was less pronounced at 0.1% supernatant concentration, consistent with a dose-dependent antioxidant effect (Fig. S4A).

Second, PBMCs from three healthy donors were co-cultured with the individual probiotic strains and stimulated with anti-CD3/anti-CD28 antibodies for 24 h. Several strains increased IL-10 release compared with stimulated PBMC controls, indicating induction of an anti-inflammatory cytokine response (Fig. 2H). Additional cytokine profiling showed reduced IL-13 and IL-17 concentrations for most strains, whereas IFN-γ was increased by a subset of strains (Fig. S4B). Together, these findings indicate that the tested probiotic strains are biologically active *in vitro* and can modulate oxidative stress and immune cytokine responses, although their effects are strain- and cytokine-dependent.

## DISCUSSION

The gut microbiome is an important regulator of immune responses and functional outcome after stroke.^19^ Experimental brain injury has been shown to alter gut microbiome composition,^20^ and stroke-induced dysbiosis can shape neuroinflammatory responses through effects on intestinal immunity.^5^ In the present study, we show that experimental and clinical ischemic stroke are associated with persistent alterations in gut microbiome composition beyond the acute phase. In mice, microbiome composition remained altered 14 days after fMCAo, while in patients, beta diversity remained distinct from controls at three months after ischemic stroke or TIA. These findings extend previous work on acute post-stroke dysbiosis and support the concept that stroke induces a sustained disturbance of host–microbiome homeostasis. Given that dysbiosis has been implicated in chronic inflammatory and vascular diseases, including hypertension,^21^ colorectal cancer, and colitis,^22^ persistent post-stroke dysbiosis may contribute to the broader systemic consequences of stroke and warrants further investigation in larger longitudinal cohorts.

The gut microbiome communicates with the brain through several pathways, including neural signaling, circulating metabolites, and immune mechanisms.^23^ Among microbial metabolites, SCFAs are of particular interest because they regulate intestinal barrier integrity, immune cell differentiation, and inflammatory responses. In our study, stroke was associated with sustained alterations in SCFA profiles. In patients, propionic and valeric acid were reduced both acutely and three months after stroke, while butyric acid showed partial recovery. In mice, experimental stroke induced compartment-specific SCFA alterations, including reduced ileal propionate and reduced total plasma SCFAs. These findings are consistent with a previous study in cynomolgus monkeys showing persistent SCFA reductions 12 months after experimental stroke.^24^ Functionally, SCFAs have been linked to improved post-stroke recovery: transplantation of SCFA-producing bacteria into aged mice improved behavioral outcome and reduced IL-17 production by γδ T cells,^25^ and SCFA supplementation improved motor recovery and cortical network remodeling after stroke.^26^ Propionate has also been shown to support blood–brain barrier integrity.^27^ Together, these data suggest that persistent disruption of microbiome-derived metabolites may influence stroke recovery and identify SCFA-related pathways as potential therapeutic targets.

Our interventional data further suggests that post-stroke microbiome alterations are modifiable. Probiotic treatment enriched administered strains in stool samples and induced measurable shifts in microbiome composition, although global alpha-diversity was not restored. Importantly, probiotic-treated patients showed greater improvement in mRS between baseline and three months, suggesting a potential effect on functional recovery. This clinical signal should be interpreted in the context of the exploratory trial size and the absence of significant effects on NIHSS or MoCA, but it provides a rationale for larger studies powered for clinical endpoints. Previous studies have reported anti-inflammatory effects of probiotic or synbiotic interventions in human cohorts,^28–32^ and our in-vitro data support biological activity of the tested strains, including reduced oxidative stress responses and modulation of PBMC cytokine release. However, inflammatory mediators and SCFA concentrations were not measured in the interventional cohort. Future studies should therefore combine clinical endpoints with longitudinal microbiome, metabolomic, and immune profiling to determine whether probiotic-induced microbiome changes translate into functional metabolic or immunological effects after stroke.

### Limitations of the study

Several limitations should be considered when interpreting these findings. First, the StrokeMicroBiomics cohort was small, which limits statistical power and may reduce the generalizability of individual taxonomic and metabolomic findings. Nevertheless, the integration of longitudinal patient sampling with experimental stroke data, metabolomic profiling, and comparison to published stroke microbiome studies supports the robustness of the main observation: persistent post-stroke microbiome and SCFA alterations.

Second, the control samples used for the StrokeMicroBiomics analysis were derived from an external cohort. Although controls were selected to match age and sex, differences in geography, sampling procedures, diet, medication exposure, and unmeasured comorbidities may have influenced microbiome composition. This is a common challenge in human microbiome studies, where age, lifestyle, disease state, and environmental factors strongly affect microbial profiles.^33^ Importantly, the longitudinal within-patient design of the stroke cohort helps to distinguish persistent post-stroke alterations from baseline inter-individual variability.

Third, the interventional cohort was exploratory in size and included predominantly mildly affected patients who were able to comply with repeated study procedures, a frequent source of selection bias in stroke trials. The study was therefore not powered to definitively establish clinical efficacy. In addition, the stool collection method used in the interventional cohort did not allow parallel fecal metabolite analysis, and inflammatory mediators were not measured longitudinally. Consequently, the mechanistic link between probiotic-induced microbiome changes, SCFA metabolism, immune modulation, and functional recovery remains to be established.

Despite these limitations, the study provides convergent experimental, observational, and interventional evidence that post-stroke microbiome alterations persist beyond the acute phase and are therapeutically modifiable. Larger prospective studies with integrated microbiome, metabolomic, immune, and clinical endpoints are warranted to validate these findings and define the mechanisms by which microbiome-targeted interventions may influence stroke recovery.

## Data Availability

All data produced in the present study are available upon reasonable request to the authors.

## RESOURCE AVAILABILITY

### Lead contact

Further information and requests for resources should be directed to and will be fulfilled by the lead contact, Arthur Liesz.

### Materials availability

This study did not generate new unique reagents.

### Data and code availability

- All original metagenomic data has been deposited at PRJEB126863 and will be made publicly available as of the date of publication.
- All original metabolomic data will be made publicly available as of the date of publication.
- Any additional information required to reanalyze the data reported in this paper is available from the lead contact upon request.

## ACKNOWLEDGMENTS

We acknowledge support from the LCSB Metabolomics, Lipidomics and Proteomics Platform (RRID:SCR_024769) in data acquisition and processing; the authors thank Christina Bauer and Kerstin Thuß-Silczak for their technical assistance; the authors want to express their gratitude to all patients and their relatives for their participation in this study.

## AUTHOR CONTRIBUTIONS

Conceptualization, P.M. and A.L.; methodology, P.M. and A.L.; Investigation, human cohorts: P.M. and C.H., murine cohorts: P.M., cell culture: M.Z. and W.M.; Software, V.A. and A.S.; Visualization: P.M., A.S. and V.A.; writing—original draft, P.M. and A.L..; writing—review & editing, P.M., C.B., P.W., and A.L.; Analysis: P.M., V.A., A.S. and C.J.; funding acquisition, A.L.; resources, P.W., C.B. and A.L.; supervision, A.L.

## DECLARATION OF INTERESTS

The authors declare receiving support for this project from Institut AllergoSan in the form of providing the probiotic/placebo compounds, a partial coverage of the project-specific shotgun sequencing costs to the sum of 10000 Euros, as well as a one-time donation in the sum of 5000 Euros for the implementation of the annual Stroke-Immunology Conference. This company had no role in the study design, conduct, data analysis, or manuscript preparation, and did not influence the decision to publish.

Furthermore, data was supplied by the manufacturer of the probiotic/placebo compounds: Winclove Probiotics. This data was used to create Figure 2 G) and H) as well as Supplemental Figure 4 A and B. Beyond this data, the company had no role in the study design, conduct, data analysis, or manuscript preparation, and did not influence the decision to publish. M.Z. and W.M. are employees of Winclove Probiotics and listed as authors due to their data contribution.

Beyond the funding and affiliations disclosed above, the authors declare no financial or personal relationships that could be perceived to influence the work reported in this paper.

## DECLARATION OF GENERATIVE AI AND AI-ASSISTED TECHNOLOGIES

During the preparation of this work, the authors did not utilize any generative AI or AI-assisted technologies.

## SUPPLEMENTAL INFORMATION

### Document S1: Figures S1-4, Tables S1-2

**Figure S1:** SCFAs in the ileum content and plasma of a cohort of C57BL6J mice are chronically altered following experimental stroke.

**Figure S2:** Expanded mass-spectrometry findings and KEGG-pathways analysis of SMB/Control cohorts

**Figure S3:** Correlation of SCFA concentrations with bacterial genus in SMB and Control patients

**Figure S4:** In-vitro probiotics testing and PRISE cohort supplementary data

**Table S1:** Characteristics of the StrokeMicroBiomics patient collective

**Table S2:** Characteristics of the PRISE patient collective

## STAR★METHODS

### KEY RESOURCES TABLE

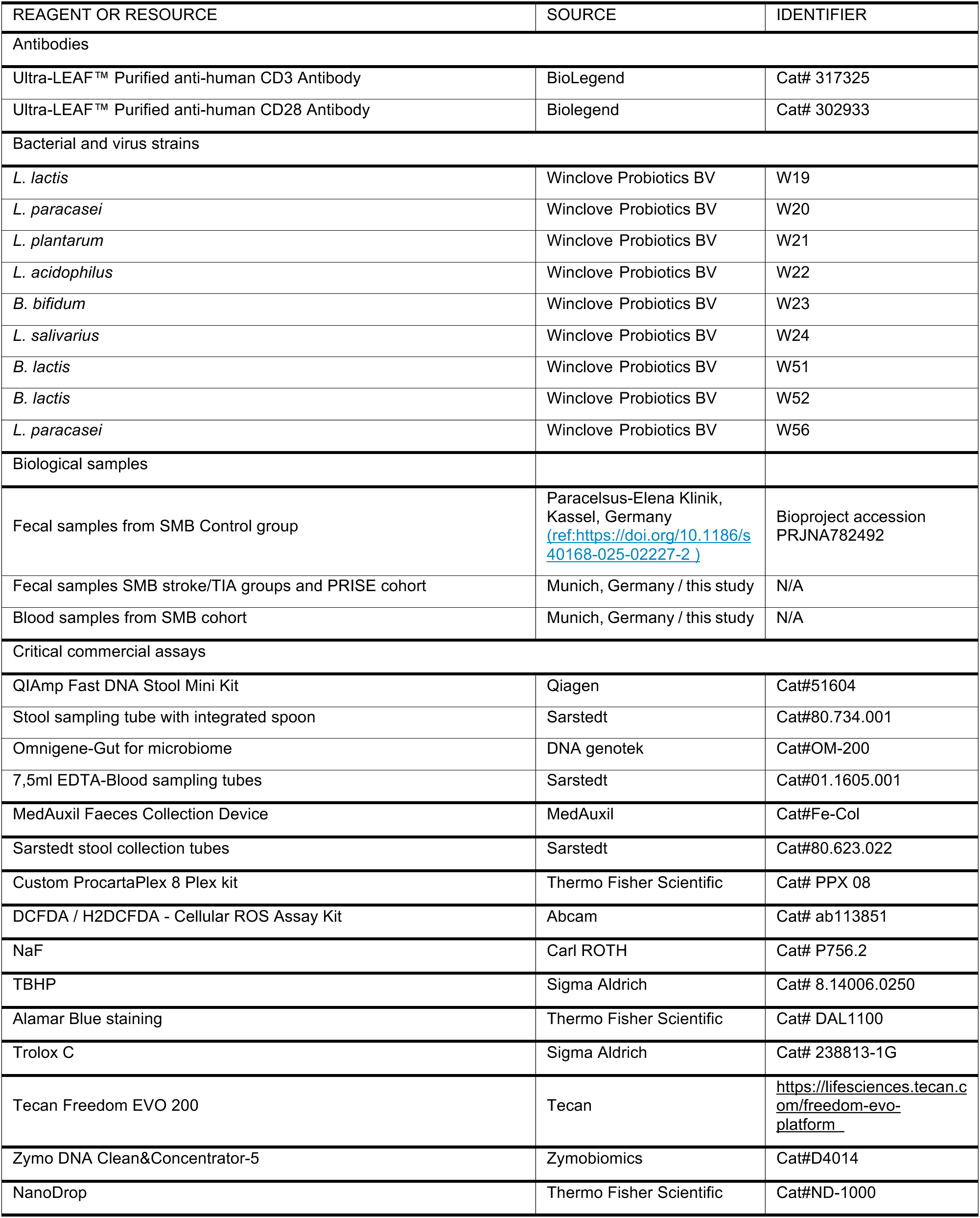
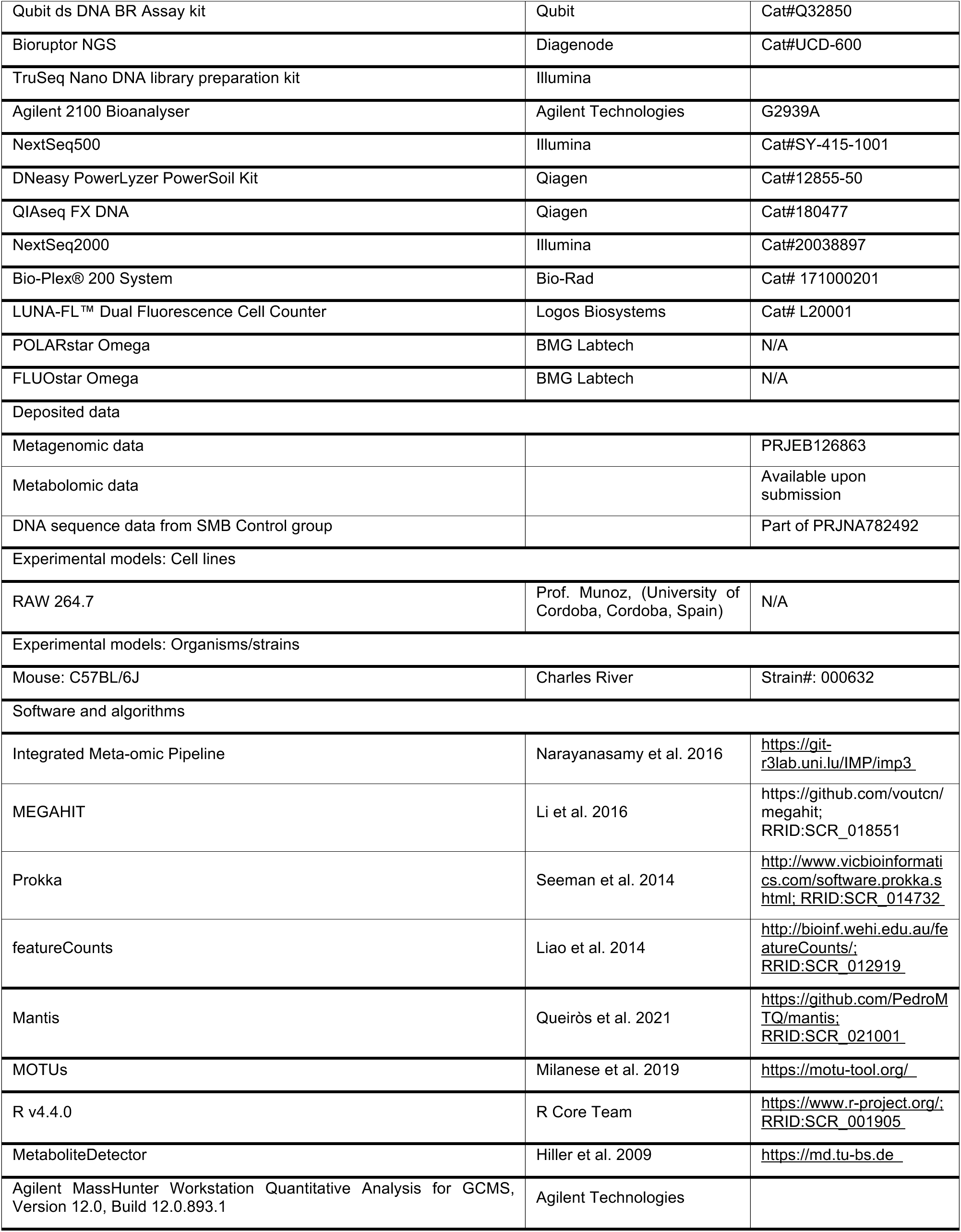
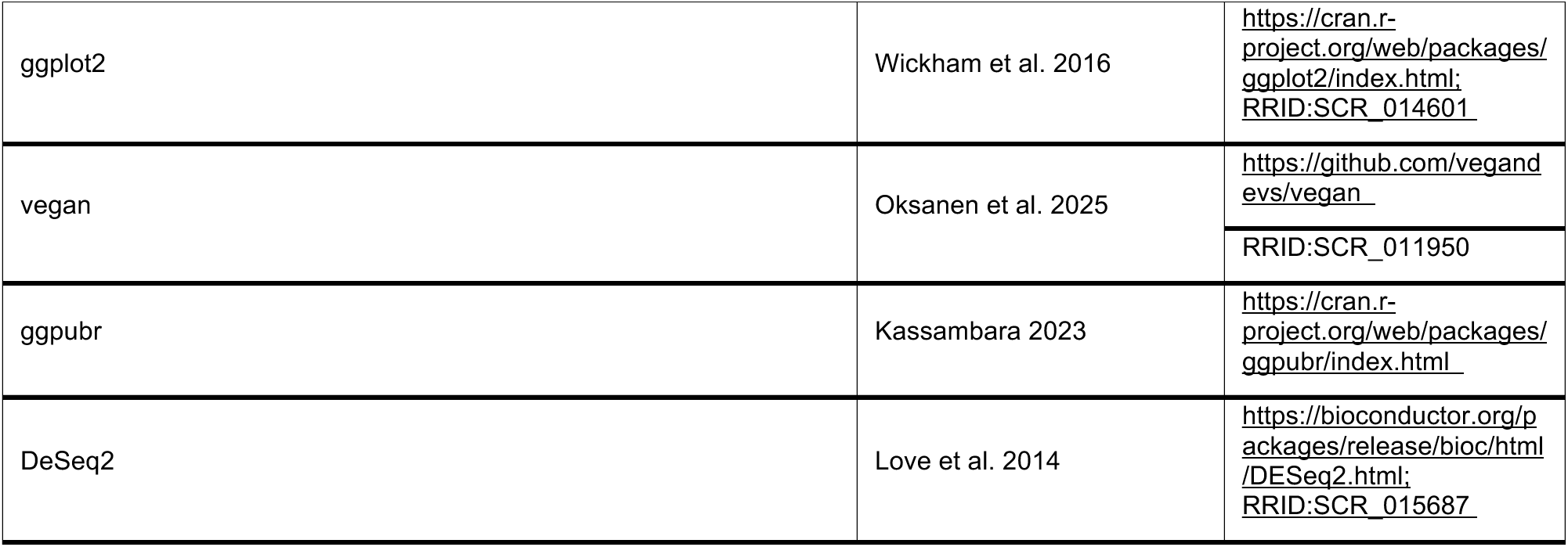

### EXPERIMENTAL MODEL AND STUDY PARTICIPANT DETAILS

Informed consent was obtained from all participants. All listed studies were performed in accordance with the Declaration of Helsinki and the current ICH GCP guidelines.

#### Clinical study populations

##### StrokeMicroBiomics cohort (observational study)

Clinical data and biological samples were acquired from 10 patients recruited to the StrokeMicroBiomics (SMB) study (NCT04315922) at the LMU Munich Hospital between 01.06.2019 and 01.02.2021. Participants in this study were required to be at least 50 years of age and have experienced either a transient ischemic attack (TIA) or stroke as confirmed by medical imaging within the last 7 days before collection of the baseline samples. Exclusion was based on the presence of one or more of the following conditions: Pregnancy, diagnosed and malignant tumor ailment, active non-stroke related immunosuppression (e.g. HIV), infection, operative procedure or antibiotics treatment within 4 weeks prior to stroke/TIA, relevant autoimmune disease (e.g., inflammatory bowel disease), chronic infectious diseases (e.g., Hepatitis C), hemorrhagic stroke or intracranial bleeding, cerebellar lesions, other neurodegenerative diseases (e.g., Parkinsońs disease or Alzheimers dementia). Sampling of stool and EDTA blood occurred at baseline and at the 3-month follow-up. A permit for house calls during the Covid-19 pandemic to collect samples and perform examinations was granted by the regional government of Bavaria. Patients who withdrew consent or could no longer participate due to severity of illness or death were excluded from analysis. Approval for this project was granted by the Ethics Committee of the Medical Faculty of the LMU Munich (121-09). Demographical and clinical characteristics of the patients are shown in Supplemental Table 1.

For the control cohort, previously published clinical and stool sampling data from 7 healthy volunteers from the DeNoPa study (DRKS00000540) was acquired.^34^ Out of the seven control subjects included in the present study, three were part of the DeNoPa cohort: a prospective, biannual follow-up study of (initially *de novo)* Parkinson’s disease (PD) patients at the Paracelsus-Elena Klinik, Kassel, Germany. Inclusion and exclusion criteria and ancillary investigations have been published previously.^35,36^ The remaining four control subjects were neurologically healthy subjects living in the same households as DeNoPa participants. The study was approved by the ethics committee of the Physician’s Board Hessen, Germany (FF 89/2008). The sample analysis for the control subjects was approved by the Comité National d’Ethique de Recherche of Luxembourg (reference no.: 140174_ND). These participants were selected to match age and sex of the stroke patients in the SMB study. Control participants (n = 7) had a median age of 71 years (IQR 60–78); 2 (29%) were male.

##### PRISE cohort (interventional trial)

Clinical data and biological samples were acquired from 22 participants recruited to the “Auswirkungen Eines Probiotischen Nahrungsergänzungsmittels Auf Das Darmmikrobiom Von Schlaganfallpatienten” (PRISE) study (NCT04954846) at the LMU Munich Hospital between 01.01.2021 and 01.03.2023.

Participants in this study were required to be at least 18 years of age and have experienced a stroke as confirmed by medical imaging within the last 7 days before collection of the baseline samples. Exclusion was based on the presence of one or more of the following conditions: chronic immunological disease, severe gastrointestinal disease including previous colectomy or an intestinal stoma, liver cirrhosis, pancreatitis, gastritis, cholecystitis or other acute gastrointestinal illness, chronic alcohol abuse, or a terminal prognosis. Patients who withdrew consent or could no longer participate due to severity of illness or death were excluded from analysis. Individuals in the probiotic group received the multispecies probiotic OMNi-BiOTiC^®^ SR-9 (Institut AllergoSan Pharma GmbH, Graz, Austria) for 90 days. Participants were instructed to take one sachet (3 g) twice daily, providing a total of 1.5 × 10¹⁰ CFU per daily dose. The probiotic contains 9 bacterial strains: *Bifidobacterium bifidum* W23, *Bifidobacterium animalis* ssp. *lactis* W51, *Bifidobacterium animalis* ssp. *lactis* W52, *Lactobacillus acidophilus* W22, *Lacticaseibacillus paracasei* W56, *Lacticaseibacillus paracasei* W20, *Lactiplantibacillus plantarum* W62, *Ligilactobacillus salivarius* W24, and *Lactococcus lactis* ssp. *lactis* W19.

Participants in the placebo group received sachets identical in appearance, consistency, and taste. The placebo formulation contained maize starch, maltodextrin, inulin, potassium chloride, magnesium sulfate, fructooligosaccharides, amylases, and manganese sulfate.

Approval for this project was granted by the Ethics Committee of the Medical Faculty of the LMU Munich (20-0942). Demographical and clinical characteristics of the patients are listed in Supplemental Table 2.

##### Peripheral blood mononuclear cells (PBMCs) from buffy coats

Buffy Coats were obtained from healthy human volunteers (NHS Blood and Transplant, Filton, Bristol) and written informed consent was obtained in each case (n=3). The collection and use of blood samples, and the research described herein, performed at Charles River Laboratories, Portishead UK, complied with relevant guidelines, institutional practices and were approved by a Research Ethics Committee (REC; IRAS ID 213880, REC reference 17/LO/0221). Donors were between 17-72 years of age and of both sexes as per NHS blood donation guidelines.

##### Animal experiments

Wild-type C57BL/6J male mice (aged 8-12 weeks) were used for the experiments. Mice were housed at the animal core facility of the Centre for Stroke and Dementia Research Munich (CSD). The holding facility for the mice was kept at regulated temperatures (22 ± 2°C), with a 12-h light–dark cycle. All mice had free access to food and water. Data were excluded from all mice that died during intervention. Animals were randomly assigned to treatment groups, and all analyses were performed by investigators blinded to group allocation. All animal experiments were performed and reported in accordance with the ARRIVE guidelines.

##### RAW 264.7 mouse macrophages cell culture

The RAW 264.7 cell line was kindly provided by Prof. Munoz, (University of Cordoba, Cordoba, Spain) to VivaCell Biotechnology GmbH, Germany, where the ROS production measurements were performed. The cells were maintained in supplemented DMEM medium (Gibco, Cat no.41966-029) containing 10% FBS (Anprotec, Cat no. AC-SM-0190) and 1% antibiotics penicillin/streptomycin (Life Technologies, Cat no.15140122) (DMEM complete medium) at 37°C in a humidified atmosphere of 5% CO2. Cells up to passage 10 were used for in vitro ROS measurements.

##### Bacterial strains and growth conditions

The probiotic strains *Lactococcus lactis subsp. lactis* W19, *Lacticaseibacillus paracasei* W20, *Lactiplantibacillus plantarum* W21, *Lactobacillus acidophilus* W22, *Bifidobacterium bifidum* W23, *Ligilactobacillus salivarius* W24, *Bifidobacterium animalis subsp. lactis* W51, *Bifidobacterium animalis subsp. lactis* W52, and *Lacticaseibacillus paracasei* W56 were obtained from the Winclove Probiotics strain collection. For cultivation, strains were first streaked onto de Man, Rogosa and Sharpe (MRS) agar plates supplemented with 0.05% (w/v) L-cysteine (Tritium M249.02) and incubated anaerobically at 37°C for 48 h until isolated colonies were obtained. A single colony was subsequently inoculated into MRS broth (110661, GranuCult®, Merck) supplemented with 0.1% (w/v) L-cysteine and cultured anaerobically at 37°C for approximately 24 h. Following incubation and OD600 quantification using BioTek Synergy H1 (Agilent), bacterial cultures were centrifuged at 1500g for 10 min at room temperature to pellet the cells. The resulting supernatants were collected and passed through a 0.2 μm sterile filter to remove any remaining bacterial cells. Cell-free supernatants were aliquoted and stored at -80°C until further use in ROS quantification assay.

### METHOD DETAILS

#### Stool sampling (SMB cohort)

Stool samples from patients were collected at baseline (within 7 days of cerebrovascular event) and at 3-month follow-up using stool sampling tubes with integrated spoon (Sarstedt). After collection, sample tubes were immediately flash-frozen in liquid nitrogen and stored at -80 °C. Shipment was performed on dry ice.

#### Blood sampling (SMB cohort)

Blood was collected in 7.5 ml Sarstedt EDTA blood sampling tube at baseline and at 3-month follow-up within 15 minutes of stool sample collection. Samples were immediately placed on ice (4 °C) and transported to the ISD laboratory upon collection. Upon arrival, EDTA blood was spun at 2000g for 10 min and the resulting plasma divided into 500 μL samples aliquots for storage at -80 °C. Shipment was performed on dry ice.

#### Stool sampling (Control cohort)

Fecal samples were collected at the clinics via a Faeces Collection Device (MedAuxil) and collection tubes (Sarstedt), as previously described.^37^ Samples were immediately flash-frozen on dry ice after collection. Samples were subsequently stored at –80 °C and shipped on dry ice.

#### Stool sampling (PRISE cohort)

Stool samples from patients were collected at 3-month follow-up using Omnigene-Gut for microbiome Om-200 (DNA genotek) collection tubes by the participants according to the manufacturers protocol. Samples from household controls were collected at the same time. Samples were kept at room temperature for up to 30 days in accordance with the manufacturer’s specifications before DNA isolation. Subsequent shipment was performed on dry ice.

#### Human shotgun metagenomic sequencing and data analysis

For the SMB stroke/TIA group’s fecal samples, DNA was extracted using the QIAmp Fast DNA Stool Mini Kit (Qiagen) following the manufacturer’s protocol. DNA quality and quantity were evaluated with an ND-1000 NanoDrop Spectrophotometer (Peqlab). The QIAseq FX DNA kit was used for library preparation. Samples were sequenced on an Illumina NextSeq2000 instrument with 2×150 bp read lengt at the Luxembourg Centre for Systems Biomedicine. For the PRISE cohort’s fecal samples, DNA was extracted using the QIAmp Fast DNA Stool Mini Kit (Qiagen) following the manufacturer’s protocol. DNA quality and quantity were evaluated with an ND-1000 NanoDrop Spectrophotometer (Peqlab). Sequencing was performed at the Gene Center Munich (Laboratory for Functional Genome Analysis) according to standard procedure.

For control subjects, extractions from fecal samples were performed according to a previously published protocol,^38^ conducted on a customized robotic system (Tecan Freedom EVO 200). After extraction, DNA was purified prior to sequencing using the Zymo DNA Clean&Concentrator-5 (D4014) kit. DNA quality and quantity were evaluated with a NanoDrop Spectrophotometer 1000 (Thermo Scientific) as well as a commercial kit from Qubit (Qubit ds DNA BR Assay kit, Q32850). Following extraction, 200-300 ng of DNA was sheared using a Bioruptor NGS (Diagenode) with 30s ON and 30s OFF for 20 cycles. Sequencing libraries were prepared using the TruSeq Nano DNA library preparation kit (Illumina) following the manufacturer’s protocol, with 350 bp average insert size. Library quality was evaluated with a Bioanalyzer (Agilent) and DNA quantified using Qubit (Invitrogen). The library was sequenced on an Illumina NextSeq500 instrument with 2×150 bp read length.

For the SMB cohort shotgun metagenome sequence data were processed using the Integrated Meta-omic Pipeline (IMP)^39^ [https://git-r3lab.uni.lu/IMP/imp3, commit identifier ecead78]. The first steps of the workflow involved trimming and quality filtering the reads and removing human reads based on mapping against a human genome reference (hg38). After pre-processing, the reads were assembled using the IMP-based iterative hybrid-assembly pipeline using MEGAHIT v. 1.0.3.^40^ Structural features such as open-reading frames (ORFs) were predicted using a modified version of Prokka,^41^ annotated with Mantis^42^ and quantified with featureCounts.^43^ Taxon abundances were estimated using mOTUs 2.5.1.^44^

For the PRISE cohort, raw sequence data was filtered and trimmed using Atria (parameters: --length-range 75:200 -q 20 -n 2 -g NO --polyG).^45^ Trimmed reads were then aligned against the human genome (GRCh38) using bowtie2 with the “--very-sensitive” flag enabled, to remove host sequences.^46^ Taxonomic profiles were generated from cleaned reads using mOTUs 2.5.1.^44^

#### Murine sample collection

Fresh stool pellets were collected at each timepoint in sterile 1.5 mL Eppendorf tubes and stored on dry ice until long-term storage at −80 °C. In preparation for internal tissue collection, the C57BL6J mice were anaesthetized using a drug cocktail consisting of ketamine (120 mg/kg) and xylazine (16 mg/kg). Blood was drawn from the right cardiac ventricle and deposited in 50 mM EDTA coated tubes (Sigma-Aldrich). Blood samples were centrifuged at 2000g for 10 minutes (4 °C) and resulting plasma stored at -80 °C. Following blood collection, mice were perfused via PBS injection to the left ventricle of the heart. The small intestine was surgically removed and ileum content placed in a 2 ml Eppendorf tube and stored at –80 °C. All shipments were performed on dry ice.

#### Murine shotgun metagenomic sequencing and data analysis

Frozen stool samples were weighed and ∼250 mg from each sample was used to extract microbial genomic DNA using the DNeasy PowerLyzer PowerSoil Kit (Qiagen) according to the manufacturer’s instructions. Concentration and purity of extracted DNA was measured on a NanoDrop ND-1000 Spectrophotometer (Thermo Fisher, United States).

For all mouse fecal samples, shotgun metagenome sequence data were processed using a custom pipeline. Raw reads were concatenated prior to quality filtering and trimming using Atria [ref: https://pmc.ncbi.nlm.nih.gov/articles/PMC10038132/], (parameters: --length-range 75:200 -q 20 -n 2 -g NO --polyG), filtering all reads with a quality score lower than 20 and shorter than 75 base pairs, and additionally trimming polyG tails. Non-microbial sequences were removed using bowtie2 with “-very-sensitive” enabled. Taxonomic abundances were estimated using mOTUs 2.5.1.^44^

#### Meta-metabolomics

Untargeted GC-MS and targeted measurements for short-chain fatty acids (SCFAs) from human fecal samples for the two cohorts were performed in separate batches, but following the same previously published protocol,^47,48^ with minor modifications. Targeted GC-MS measurements of murine plasma and ileum content samples for SCFAs were performed on an Agilent 8890 GC – 5977B MS following the same protocol.

For fecal SCFA extraction, briefly, samples were homogenized in a water–internal standard solution containing 2-ethylbutyric acid (2 mmol/L) at a ratio of 500 µL per 50 mg of fecal matter. The homogenates were acidified and incubated, followed by extraction with diethyl ether. The resulting extracts were derivatized using N-tert-butyldimethylsilyl-N-methyltrifluoroacetamide (MTBSTFA) prior to GC-MS analysis; detailed measurement parameters have been described previously. ^47^

Plasma SCFA extraction was performed as follows. Briefly, 50 µL of plasma were mixed with 50 µL of an aqueous spiking solution containing 2 mmol/L 2-ethylbutyric acid (internal standard) and 1 mmol/L Volatile Free Acid Mix (VFAM; Sigma Aldrich) using a standard addition approach, yielding final pre-filtration concentrations of 1 mmol/L internal standard and 0.5 mmol/L VFAM. After vortexing, 100 µL were transferred to pre-rinsed Amicon Ultra 0.5 Ultracel 10K centrifugal filter units and centrifuged (14,000 × g, 15 min, 21 °C). From the filtrate, 60 µL were acidified with 3 µL 37% HCl, mixed in a ThermoMixer (15 °C, 2,000 rpm, 15 min), extracted twice with diethyl ether (300 µL each; 15 °C, 2,000 rpm, 15 and 5 min, respectively), and the resulting organic phases (2 × 250 µL) were combined in 2 mL tubes. Extracts were vortexed, aliquoted in triplicate (150 µL) into GC vials with microinserts, derivatized with 15 µL N-methyl-N-(tert-butyldimethylsilyl)trifluoroacetamide containing 1% tert-butyldimethylchlorosilane (MTBSTFA) for ≥2 h at room temperature, and analyzed by GC-MS under the same conditions as described for fecal samples.

GC-MS chromatograms (untargeted) were processed using MetaboliteDetector, v3.220190704.^49^ Compounds were initially annotated by retention time and mass spectrum using an in-house mass spectral library. Internal standards were added at the same concentration to every medium sample to correct for uncontrolled sample losses and analyte degradation during metabolite extraction. The data was normalized by using the response ratio of the integrated peak area of the analyte and the integrated peak area of the internal standard.

For untargeted LC-MS, metabolite extraction from plasma was performed using a chilled (4 °C) internal standard (IS) mix containing the following compounds at their final working concentrations: [UL-¹³C₅]ribitol, pentanedioic-d₆ acid, sodium-D-3-hydroxybutyrate-¹³C₄, and D-mannose-¹³C₆ at 2 µg/mL; tridecanoic-d₂₅ acid, 6-chloropurine riboside, 4-chloro-DL-phenylalanine, Nε-trifluoroacetyl-L-lysine, and thionicotinamide adenine dinucleotide at 10 µg/mL. Internal standards were obtained from Omicron Biochemicals, C/D/N Isotopes, Eurisotop, and Sigma-Aldrich. Plasma samples were thawed on ice and centrifuged at >16,000 ×g for 3 minutes at 4 °C. A total of 75 µL of supernatant was mixed with 30 µL of the chilled IS mix and vortexed thoroughly. For protein precipitation, 320 µL of cold methanol (4 °C) was dispensed into a microcentrifuge tube, and 80 µL of the plasma/IS mixture was rapidly added to ensure immediate mixing. The samples were vortexed, incubated for 15 minutes at 4 °C at 2000 rpm (Eppendorf ThermoMixer), and centrifuged at >16,000 ×g for 5 minutes at 4 °C. For LC-MS analysis, 150 µL of the supernatant was transferred to a new tube and evaporated in a SpeedVac (Labconco) at –4 °C overnight. Samples were reconstituted in 50 µL of 80% acetonitrile in MilliQ water. Prior to analysis, samples were filtered using 0.2 µm PHENEX-RC syringe filters (Phenomenex).

Fecal samples were processed as follows: 50 mg of feces (dry weight) was suspended in 500 µL MilliQ water (1:10 w/v) in 2.0 mL tubes containing 1.4 mm ceramic beads. Homogenization was performed at 6000 rpm for 30 s using the Precellys24 system. The homogenate was incubated (2000 rpm, 5 min, 4 °C, Eppendorf ThermoMixer) and centrifuged (>16,000 ×g, 5 min, 4 °C). The resulting supernatant (fecal water) was collected and kept on ice until further processing. For extraction, 100 µL of fecal water was mixed with 40 µL of a pre-chilled internal standard mix prepared as described above. To precipitate proteins, 320 µL cold methanol (−20 °C) was added to 80 µL of the IS-fortified fecal water. After thorough mixing, samples were incubated (2000 rpm, 5 min, 4 °C) and centrifuged (>16,000 ×g, 5 min, 4 °C). For liquid-liquid extraction, 280 µL chloroform and 180 µL MilliQ water were added to 350 µL of the supernatant to achieve a biphasic system (chloroform/methanol/water, 1:1:0.9, v/v/v). Following vortexing, samples were incubated (2000 rpm, 10 min, 4 °C) and centrifuged again (>16,000 ×g, 5 min, 4 °C) to induce phase separation. The 200 µL of the polar phase was filtered (PHENEX-RC, 0.2 µm), transferred to fresh tubes, and dried as described above. Samples were reconstituted in 80 µL of 80% acetonitrile prior to LC-MS injection.

Metabolite analysis was conducted using a Thermo Vanquish Flex Quaternary LC system coupled to a Thermo Q Exactive HF Orbitrap mass spectrometer. Chromatographic separation was performed on a SeQuant ZIC-pHILIC polymer column (5 µm, 150 × 2.1 mm) with a matching guard column (20 × 2.1 mm), maintained at 45 °C. The flow rate was 0.2 mL/min, using 20 mmol/L ammonium carbonate in water (pH 9.2, Eluent A) and acetonitrile (Eluent B) as mobile phases. The gradient program was as follows: 0–3 min, 80% B; 3–18 min, linear decrease to 20% B; 18–19 min, hold at 20% B; 19–20 min, return to 80% B; 20–25.5 min, re-equilibration at 80% B (0.2 mL/min); 25.5–29.5 min, continued re-equilibration at 80% B with increased flow rate (0.4 mL/min). The injection volume was 5 µL. Total run time: 30 min.

Mass spectrometry was performed with electrospray ionization in polarity switching mode (+ESI/−ESI). Source parameters were: sheath gas flow rate 25, auxiliary gas 15, sweep gas 0, spray voltage 4.5 kV (+) / 3.5 kV (−), capillary temperature 325 °C, S-lens RF level 50, and auxiliary gas heater temperature 50 °C. Full-scan MS spectra were acquired at a resolution of 60,000 (m/z 75–1000), with an AGC target of 1e6 and maximum injection time of 240 ms.

#### Transient proximal cerebral artery occlusion model

Transient intraluminal occlusion of the middle cerebral artery (MCA) was performed as previously described.^50^ Briefly, mice were anaesthetized with isoflurane delivered in 100% O2. A midline neck incision was made and the common carotid artery and left external carotid artery were isolated and ligated; a 2-mm silicon-coated filament was introduced via a small incision in the external carotid artery and advanced towards the internal carotid artery, thereby occluding the MCA. MCA was occluded for 60 min for the transient MCAo stroke model. After occlusion, the animals were re-anesthetized, and the filament was removed. MCA occlusion and reperfusion were confirmed by the corresponding decrease or increase in the blood flow, respectively, measured by a laser Doppler probe affixed to the skull above the MCA territory (decrease in the laser Doppler flow signal >80% of baseline value and increase in the laser Doppler flow signal>80% of baseline value before reperfusion). After recovery, the mice were kept in their home cage with facilitated access to water and food. Body temperature was maintained at 37°C throughout surgery using a feedback-controlled heating pad. The overall exclusion rate of stroke mice was approximately 50 %. Exclusion criteria: 1. Insufficient MCA occlusion (a reduction in blood flow to >20% of the baseline value); 2. Insufficient MCA reperfusion (an increase in blood flow of >80% of the baseline value before removing the filament); 3. Death before 14-day follow-up. 4. Severity of illness precluded possibility of sample collection.

#### Peripheral blood mononuclear cell (PBMC) isolation

Peripheral blood mononuclear cells (PBMCs) were isolated from human buffy coats (typically 50–60 mL) obtained from NHS Blood and Transplant (NHSBT) by density gradient centrifugation. Buffy coats were diluted with wash buffer consisting of Hank’s Balanced Salt Solution (HBSS; Gibco, 11500476) supplemented with 20 mM HEPES (Sigma Aldrich, H0887) to a final volume of 120 mL. Aliquots of 30 mL diluted buffy coat were carefully layered over 15 mL Histopaque (Sigma Aldrich, 10771) in 50 mL conical tubes and centrifuged (Heraeus Megafuge 40R, Thermo Scientific) at 500 g for 30 min at room temperature with the brake off. Following centrifugation, the PBMC layer at the plasma–Histopaque interface was collected using a Pasteur pipette and transferred to fresh tubes. Samples were brought to a final volume of 45 mL with wash buffer and centrifuged at 300 g for 10 min at room temperature. The supernatant was discarded, and cells were washed twice more in wash buffer, centrifuging at 200 g for 20 min and subsequently at 300 g for 10 min, respectively. Cell pellets were resuspended in 5 mL ACK red blood cell lysis buffer (Gibco, 11509876) and incubated for 5 min at room temperature. Lysis was quenched by adding wash buffer to a final volume of 45 mL, followed by centrifugation at 300 g for 10 min at room temperature. The supernatant was discarded, and the cell pellet was resuspended in wash buffer, passed through a 70 µm cell strainer (Corning, 431751), and counted using a Luna FL Dual Fluorescence Cell Counter (Logos Biosystems).

#### PBMC culture with bacteria

PBMCs were isolated and seeded at 5e5 cells per well (200 µL final volume) into flat bottom 96 well plates (Corning, 3595) in antibiotic-free RPMI complete medium (Gibco, 31870). T cell activation was induced using immobilized anti CD3 antibody (0.01 µg/well) (BioLegend, 317325), coated overnight at 4 °C, together with soluble anti CD28 antibody (1 µg/well) (BioLegend, 302933). Each experimental condition was performed in duplicate. Following stimulation, PBMCs were incubated for 24h with selected bacterial strains at a multiplicity of infection (MOI) of 1. Bacterial strains were prepared from previously stored glycerol stocks provided by Winclove BV (Amsterdam, The Netherlands). Frozen stocks were thawed and centrifuged at 4,000 rpm for 10 min (Heraeus Megafuge 40R) to pellet bacteria. Pellets were washed once in RPMI complete medium supplemented with 10% human AB serum (BioIVT, H4522), 4 mM L glutamine (Gibco, 25030 024), and 20 mM HEPES (Sigma Aldrich, H0887) to remove glycerol, debris, and non-viable cells. Bacteria were then resuspended, diluted to the appropriate concentration, and added to PBMC cultures at the specified MOI. To confirm bacterial viability and dosing, bacterial suspensions were plated on de Man– Rogosa–Sharpe (MRS) agar supplemented with 0.1% cysteine (Millipore, 69964; Sigma Aldrich, C8755). Back plating of inoculated bacterial suspensions was additionally performed to assess strain recovery. Unstimulated and stimulated control conditions were included in each experiment.

#### Cytokine measurement by Luminex

Cytokine concentrations in PBMC culture supernatants were quantified using custom ProcartaPlex multiplex immunoassays (Thermo Fisher Scientific). PBMC supernatants were analyzed using a custom ProcartaPlex 8 Plex kit (PPX 08), according to the manufacturer’s instructions. Samples were acquired on a Bio Plex 200 System (Bio Rad) with a minimum bead count threshold of 50 beads per analyte and a sample timeout of 60 s. Doublet discrimination (DD) gate overrides were set to 7,000–25,000 for PBMC samples. Standard curves were generated using a 5-parameter logistic (5PL) curve-fitting model in Bio-Plex Manager software (Version 6.2, Build 175). Values below the lower limit of quantification (LLOQ) or above the upper limit of quantification (ULOQ) were plotted at the functional LLOQ or ULOQ, respectively, calculated as the plate-specific LLOQ/ULOQ multiplied by the appropriate dilution factor for each sample.

#### Cytotoxicity assays (accompanying ROS production assay)

RAW 264.7 cells were seeded at a density of 1×104 cells/well, 100µl in total in 96-well plates (Falcon,Cat no.353072) and incubated overnight in DMEM medium (Gibco, Cat no.41966-029) supplemented with 10% FCS (Anprotec, Cat no. AC-SM-0190) 1% Antibiotics (Life Technologies,Cat no.15140122) at 37°C in a humidified atmosphere of 5% CO2. Then, cell cultures were stimulated with selected concentrations of the bacterial supernatants (4 doses, n=4) alone for 24 h. Cell viability was determined by Alamar Blue staining (cat no DAL1100, Life Technologies) using 100, 250 and 500µg/ml NaF (cat no. P756.1, Roth) as toxic control. AlamarBlue is a redox indicator that yields a colorimetric change and a fluorescent signal in a response to a metabolic activity. The fluorescence reaction is determined by an Microplate reader (PolarStar, BMG) measuring the intensity of the red colour as marker of the increased AlamarBlue reaction (570 nm with 600 nm as reference wavelength). This colorimetric determination of colour intensity was expressed as arbitrary units. The decrease of metabolism/cell viability was expressed in comparison to untreated control cells. Effects significantly higher than a 25% decrease in this assay were concluded to be due to cell toxic effects or slowing down cell metabolism m (<75% viability).

#### ROS production

The intracellular accumulation of ROS was detected by fluorimetry using 2’,7-’dihydrofluoresceindiacetate (DCFH-DA) (Cat. no. ab113851: DCFDA / H2DCFDA - Cellular ROS Assay Kit). DCFH-DA enters cells passively and is deacetylated by esterase to nonfluorescent DCFH. DCFH reacts with ROS to form DCF, the fluorescent product. Cells were seeded into 96-well culture plates (1 × 104 cells/well) in DMEM (Gibco, Cat no.41966-029) supplemented with 10% FBS (Anprotec, Cat no. AC-SM-0190); culture medium was renewed when the cells reached 80% confluence and then cells were prestimulated with the ingredients for 30 minutes before intracellular ROS was induced by the ROS inducer tertiary butyl hydroperoxide (TBHP, 200 µM) (Cat no.8.14006.0250, Sigma-Aldrich). Trolox C (25 µM) (238813-1G, Sigma-Aldrich) was used as antioxidant control. Intracellular ROS was detected by using a ROS detection kit (Abcam) according to the manufacturers protocol (1 h stimulation or other time points). DCF fluorescence was detected using a fluorescence reader (FLUOStar - BMG LabTechnologies Instruments Offenburg, Germany) with excitation/emission wavelengths of 488 nm/525 nm. Values after 4 h of exposure, data were normalized to the value of the control with 200 µM of THBP set as 100%.

### QUANTIFICATION AND STATISTICAL ANALYSIS

All statistical analyses, except the *in vitro* probiotic experiments, were performed in R (4.4.0) [ref: R Core Team [2024]]. Alpha (Richness) and beta diversity (Bray-Curtis distance) were calculated in R, using phyloseq (v1.52.0) and vegan (2.6.6). Differential abundance analysis was performed using the R package DESeq2 (v1.44.0). Random effects meta-analysis of Prevotella was performed using the R package metafor (v4.6.0), utilizing random effects meta-analysis on pooled effect sizes. The probiotic in-vitro experiments were analyzed in python v3.12 using the python package statsmodels version 0.15.0. Specific statistical tests are described in detail in the respective figure legends.

### ADDITIONAL RESOURCES

The clinical trial registry links for the SMB and PRISE cohorts can be accessed via their respective URLs: SMB: https://clinicaltrials.gov/study/NCT04315922

PRISE: https://ichgcp.net/de/clinical-trials-registry/NCT04954846

---

**Supplementary Figure 1:**
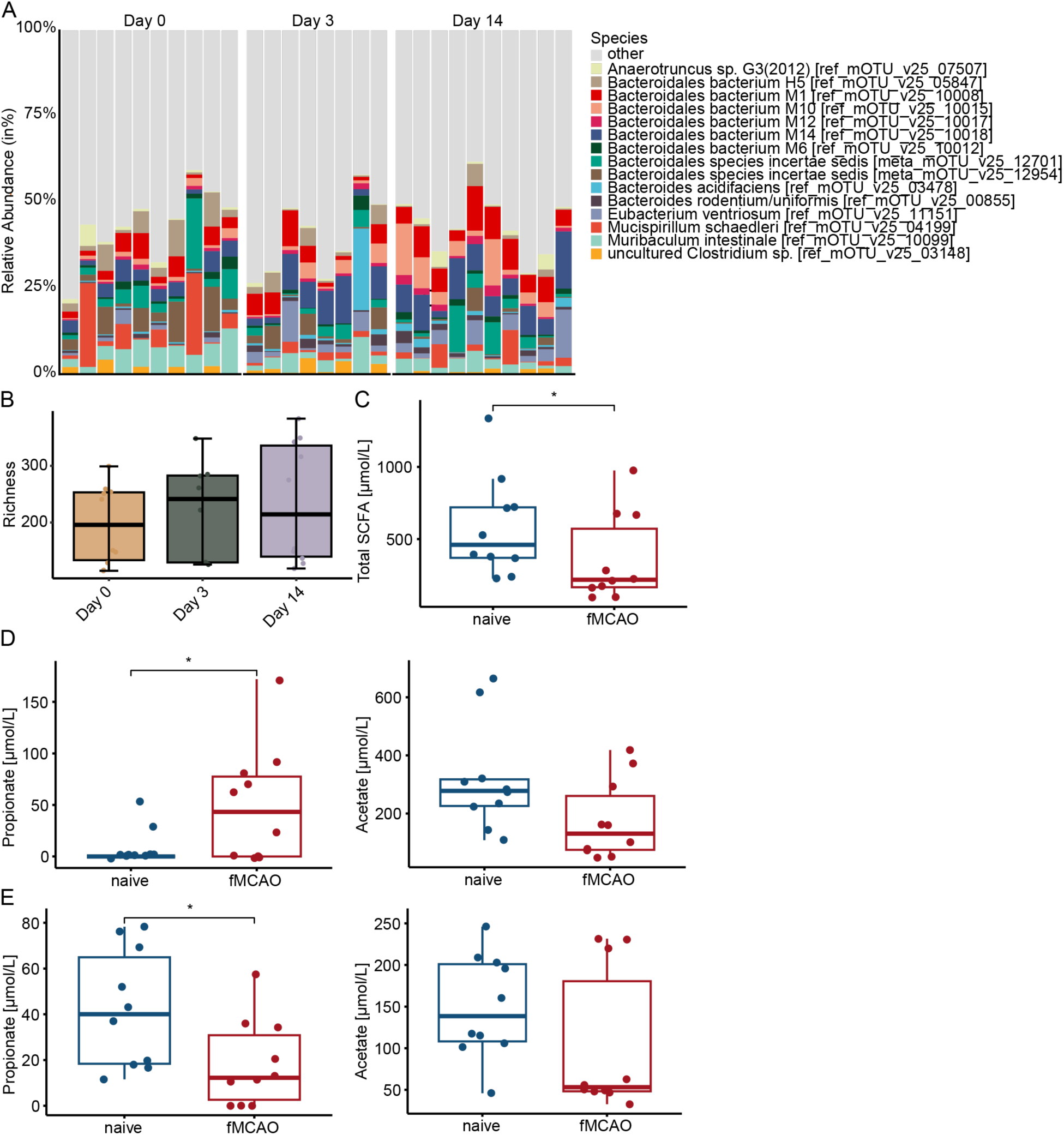
SCFAs in the ileum content and plasma of a cohort of C57BL6J mice are chronically altered following experimental stroke. (A) Relative abundance of bacterial species measured in the stool of fMCAo mice at different timepoints (N= 8-10 per time point). (B) Box-whisker plot of observed species richness measured between all time points (t-test; n = 10 per time point). (C) Box-whisker plot of total SCFA concentration (in µmol/L) measured in the plasma of surgery-naive or 14 days post-fMCAo-surgery mice (t-test; N = 10 per group; p = 0.043). (D) Box-whisker plots of concentration (in µmol/L) of propionate (left panel; t-test; N = 10 per group; p = 0.045) and acetate (right panel; t-test; N = 10 per group; p = 0.075) measured in the plasma of surgery-naive or 14 days post-fMCAo-surgery mice. (E) Box-whisker plots of concentration (in µmol/L) of propionate (left panel; t-test; N = 10 per group; p = 0.026) and acetate (right panel; t-test; N = 10 per group; p = 0.22) measured in the ileum content of surgery-naive or 14 days post-fMCAo-surgery mice. (* denotes p ≤ 0.05; ** denotes p ≤ 0.005)

**Supplementary Figure 2:**
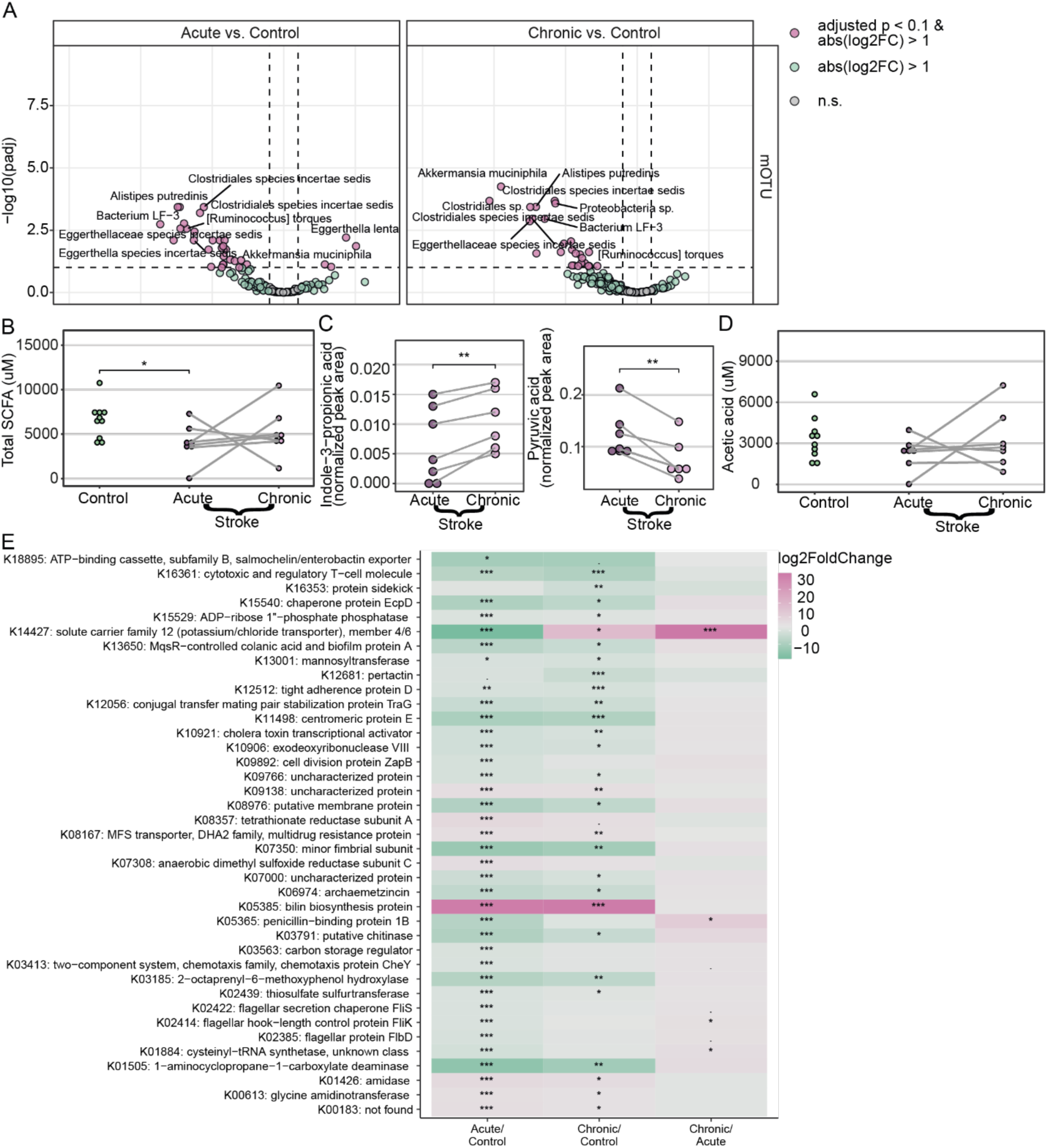
Expanded mass-spectrometry findings and KEGG-pathways analysis of SMB/Control cohorts (A) Volcano plots of differentially abundant bacterial mOTUs (DESeq2; N = 6-7; Only taxa present in ≥ 10 samples were included). (B) Dot plot showing abundance (in uM) of all SCFAs in the control, acute stroke and chronic stroke patient groups (t-test; N = 7-10). (C) Dot plot showing abundance (normalized peak area) of Indole−3−propionic acid (left panel; t-test; N = 6-7; p = 0.003) and pyruvic acid (right panel; t-test; N = 6-7; p = 0.003) in the acute and chronic stroke patient groups. (D) Dot plot showing abundance (in uM) of acetic acid in the control, acute stroke and chronic stroke patient groups (t-test; N = 7-10). (E) Heatmap showing pathway analysis of differentially regulated KEGG-orthologs between the control, acute stroke and chronic stroke groups. (* denotes p ≤ 0.05; ** denotes p ≤ 0.005; *** denotes p ≤ 0.0005)

**Supplementary Figure 3:**
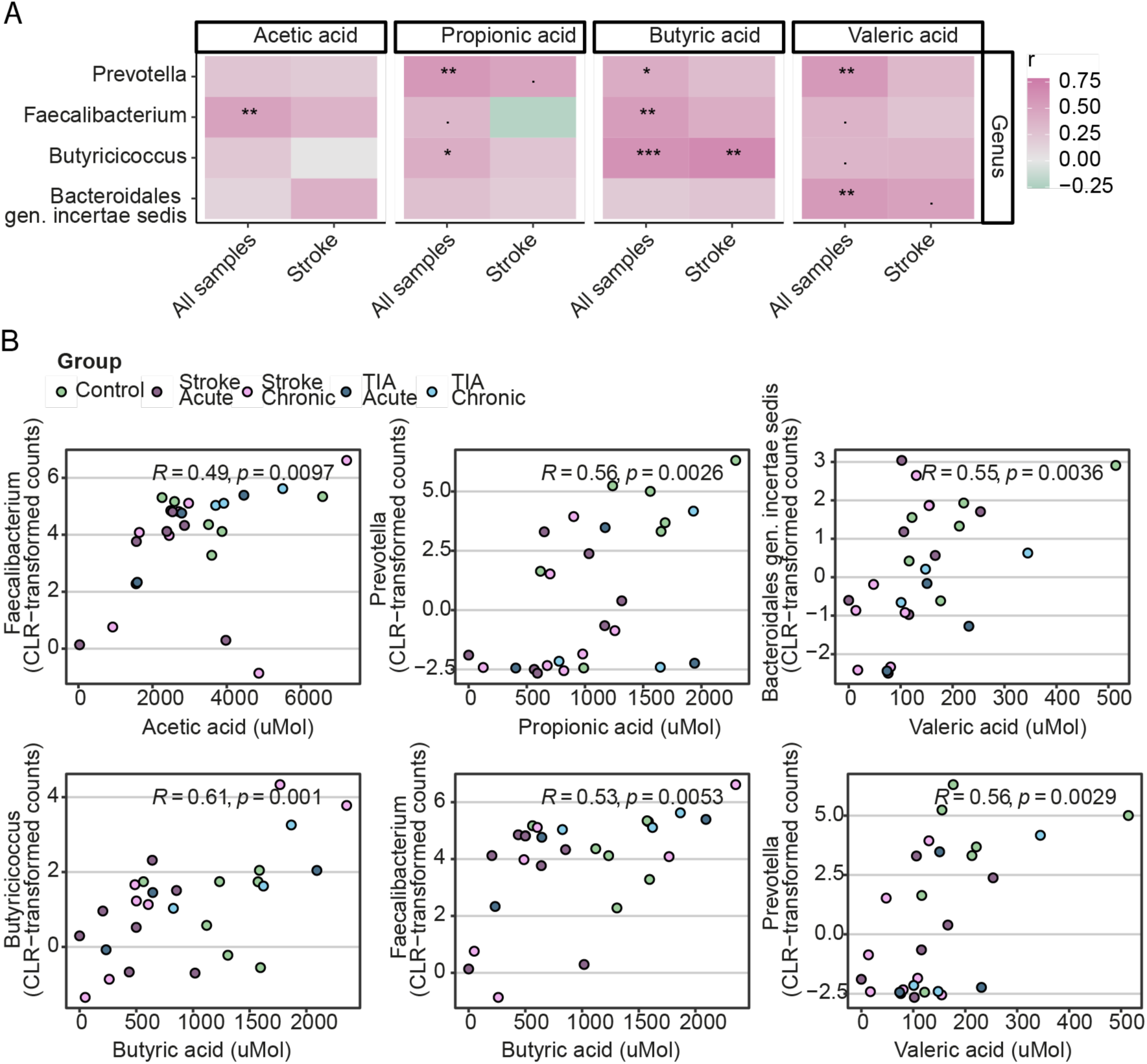
Correlation of SCFA concentrations with bacterial genus in SMB and Control patients (A) Heatmap showing the correlation (expressed with the spearman coefficient r) between bacterial genera and the concentrations of SCFAs in all samples (N = 17) and stroke samples (N = 7). (B) Correlation plots of stroke-patient-only correlations between an SCFA (in uMol) and bacterial genus (in CLR transformed counts) pair (R = spearman coefficient; pairs with R > 0 and p < 0.01 were included). (* denotes p ≤ 0.05; ** denotes p ≤ 0.005; *** denotes p ≤ 0.0005)

**Supplementary Figure 4:**
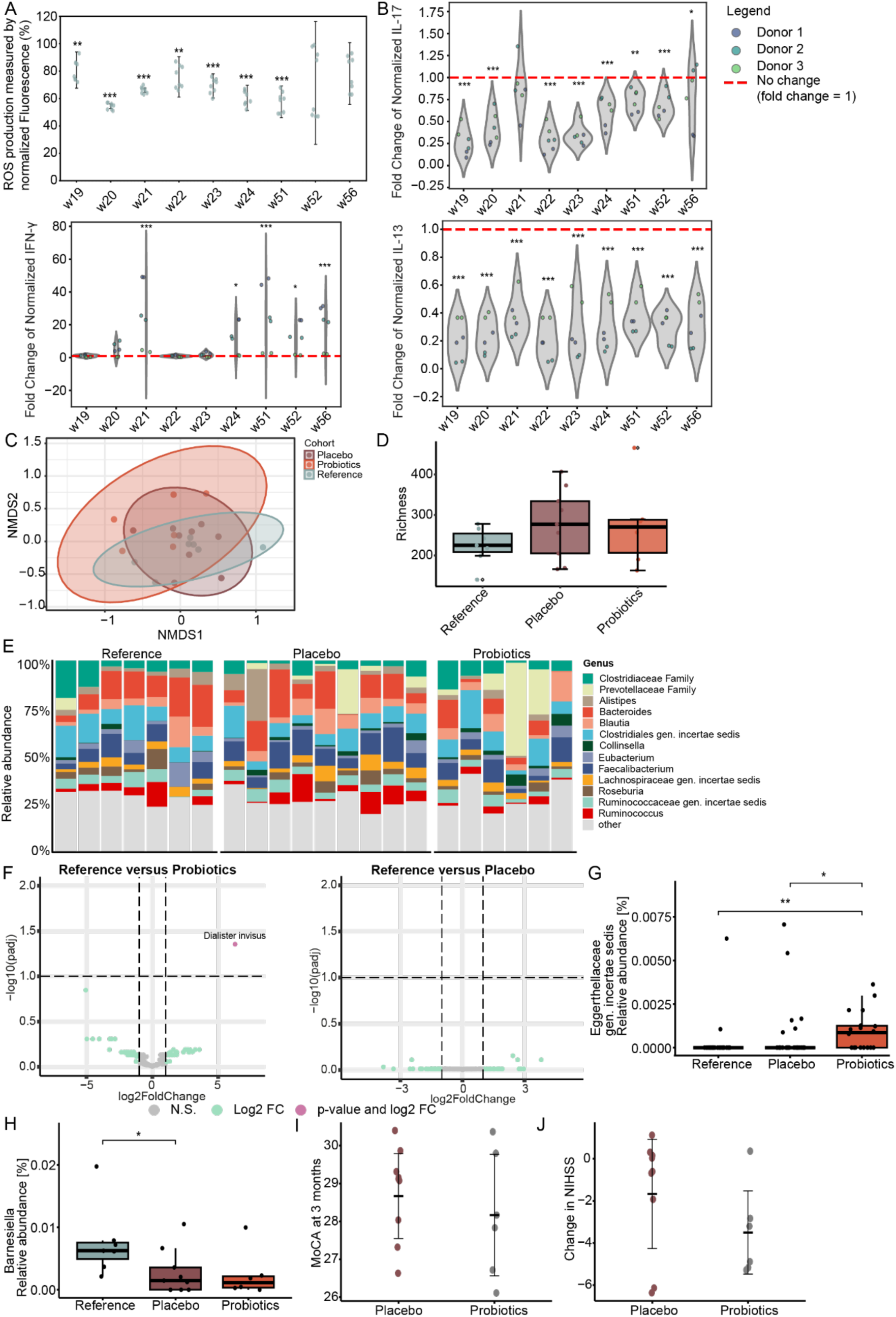
In-vitro probiotics testing and PRISE cohort supplementary data (A) Scatterplot of human macrophage cell cultures stimulated with 200 um tertiary butyl hydroperoxide and 0.1% supernatant from a bacterial strain. Radical oxygen species levels were determined via DCF fluorescence at excitation/emission wavelengths of 488 nm/525 nm. The relative changes in fluorescence were normalized to macrophages only stimulated with 200 um TBHP. (Linear mixed effects model; N = 8 per group) (B) Violin plot of fluorescence intensity fold changes of IL-17 (Top right panel), IFN-γ (bottom left panel) and IL-13 (bottom right panel) in supernatant of human PBMCs from 3 donors cocultured with probiotic strains. PBMCs were cocultured 1:1 with 9 different bacterial strains and stimulated with Anti-CD3 and Anti-CD28 antibodies for 24 hours before Luminex measurements. PMBCs stimulated without coculture were used to normalize the measurements on a per-donor basis (Linear mixed effects model; N = 6 per group). (C) Non-metric multidimensional scaling (NMDS) analysis using Bray-Curtis indices from sequenced stool samples (Adonis; N = 6-9 per group; R² = 0.11; p = 0.126). (D) Box-whisker plot of observed species richness measured between all patient groups (t-test; n = 6-9). (E) Bar graph of relative abundance (in %) of bacterial genera measured in stool of PRISE participants (F) Volcano plots of differential abundance of mOTUs between household control/probiotic groups (left panel) and household control/placebo groups (DESeq2; N = 6-9). (G) Box-whisker plot of relative abundance of Eggerthellaceae gen. incertae sedis (t-test; N = 6-9). (H) Box-whisker plot of relative abundance of genus Barnesiella (t-test; N = 6-9). (I) Dot-whisker plot of MOCA value at 3 months in the treatment groups (wilcoxon; N = 6-9; p = 0.59). (J) Dot-whisker plot of change in NIHSS between baseline and 3 months in the treatment groups (wilcoxon; N = 6 - 9; p = 0.23). (* denotes p ≤ 0.05; ** denotes p ≤ 0.005)

**Supplementary Table 1:** Characteristics of the StrokeMicroBiomics patient collective.

| Characteristic | TIA (n = 3) | Mild stroke (n = 4) | Severe stroke (n = 3) | Controls (n = 7) |
| --- | --- | --- | --- | --- |
| Age, years, median (IQR) | 60 (58–65) | 67 (60–73) | 85 (70–86) | 71 (60–78) |
| Male sex, n (%) | 2 (67) | 2 (50) | 0 (0) | 2 (29) |
| NIHSS at baseline, median (IQR) | 0 (0–2) | 2 (1–2) | 13 (13–16) | – |
| NIHSS at 3 months, median (IQR) | 0 (0–0) | 1.5 (1–2) | 6 (4–10) | – |
| mRS at baseline, median (IQR) | 0 (0–0) | 2 (1–2) | 4 (4–4) | – |
| mRS at 3 months, median (IQR) | 0 (0–0) | 1 (1–1) | 4 (3–4) | – |
| Infarct volume, mL, median (IQR) | 0.1 (0.0–0.1) | 7.2 (1.1–13.4) | 47.4 (32.4–81.7) | – |
| IV thrombolysis, n (%) | 0 (0) | 0 (0) | 2 (67) | – |
| Mechanical thrombectomy, n (%) | 0 (0) | 1 (25) | 2 (67) | – |
| Antibiotics before sampling, n (%) | 0 (0) | 0 (0) | 1 (33) | 0 (0) |

IQR, interquartile range; MCA, middle cerebral artery; mRS, modified Rankin Scale; NIHSS, National Institutes of Health Stroke Scale; TIA, transient ischemic attack. Control participants were drawn from an external cohort and matched to the stroke group on age and sex. TIA participants were not included in statistical comparisons.

**Supplementary Table 2:** Characteristics of the PRISE patient collective.

| Characteristic | Placebo (n = 9) | Probiotic (n = 6) | Household controls (n = 7) |
| --- | --- | --- | --- |
| Age, years, median (IQR) | 64 (57–79) | 62 (58–70) | 76 (49–79) |
| Male sex, n (%) | 7 (78) | 5 (83) | 1 (14) |
| NIHSS at baseline, median (IQR) | 2 (1–3) | 4 (4–5) | – |
| NIHSS at 3 months, median (IQR) | 0 (0–1) | 0.5 (0–1) | – |
| mRS at baseline, median (IQR) | 1 (0–2) | 2 (2–3) | – |
| mRS at 3 months, median (IQR) | 1 (1–1) | 1 (0–1) | – |
| MoCA at 3 months, median (IQR) | 29 (28–29) | 28 (27–30) | – |
| BDI at 3 months, median (IQR) | 5 (2–7) | 2.5 (1–4) | – |
| GSRS-IBS at 3 months, median (IQR) | 14 (13–18) | 15.5 (14–18) | – |
| Infarct volume, mL, median (IQR) | 0.7 (0.2–9.4) | 10.5 (7.7–24.8) | – |
| IV thrombolysis, n (%) | 1 (11) | 3 (50) | – |
| Mechanical thrombectomy, n (%) | 1 (11) | 5 (83) | – |
| Antibiotics before sampling, n (%) | 1 (11) | 0 (0) | 0 (0) |

BDI, Beck Depression Inventory; GSRS-IBS, Gastrointestinal Symptom Rating Scale for Irritable Bowel Syndrome; IQR, interquartile range; MCA, middle cerebral artery; MoCA, Montreal Cognitive Assessment; mRS, modified Rankin Scale; NIHSS, National Institutes of Health Stroke Scale. Household controls were cohabitants of trial participants and were not assessed clinically.

## Notes

### Clinical Trial

NCT04954846

### Author Declarations

Approval for this project was granted by the Ethics Committee of the Medical Faculty of the LMU Munich (121-09). Approval for this project was granted by the Ethics Committee of the Medical Faculty of the LMU Munich (20-0942) Buffy Coats were obtained from healthy human volunteers (NHS Blood and Transplant, Filton, Bristol) and written informed consent was obtained in each case (n=3). The collection and use of blood samples, and the research described herein, performed at Charles River Laboratories, Portishead UK, complied with relevant guidelines, institutional practices and were approved by a Research Ethics Committee (REC; IRAS ID 213880, REC reference 17/LO/0221). The study was approved by the ethics committee of the Physician's Board Hessen, Germany (FF 89/2008). The sample analysis for the control subjects was approved by the Comité National d'Ethique de Recherche of Luxembourg (reference no.: 140174_ND).

